# Population-Based Study of anti-SARS-CoV-2, Social Distancing and Government Responses in Joinville, Brazil

**DOI:** 10.1101/2021.02.08.21251009

**Authors:** Henrique Diegoli, Vivianne Samara Conzatti, Suleimy Cristina Mazin, Juliana Safanelli, Louise Domenguini Chiaradia Dellatorre, Keli Bett, Roselaine Elisa Radtke, Giulia Murillo Wollmann, Helbert do Nascimento Lima, Paulo Henrique Condeixa de França, Jean Rodrigues da Silva

## Abstract

**Background:** The city of Joinville had been mildly affected by the COVID-19 pandemic until June 2020. This study aimed to longitudinally assess the prevalence of exposure to the virus and social distancing practices in the local population.

**Methods:** A randomized selection of households stratified by region was created. From June 15 to August 7, 2020, a dweller was randomized in each household, answered a questionnaire, and performed a test for the detection of SARS-CoV-2 antibodies. The prevalence of positive tests was calculated for each week and adjusted for the test’s sensitivity and specificity.

**Results:** The adjusted proportion of positive results increased from 1.4% in the first week (margin of error [ME] 0% to 2.87%) to 13.38% in the eighth week (ME 10.22% to 16.54%). Among the 213 participants that tested positive, 55 (25.82%) were asymptomatic. Only 37 (17.37%) sought medical consultation for any symptom. Among the 77 (36.15%) that were leaving home to work or study, only 18 (23.38%) stopped due to any symptom. The proportion that referred going to bars, restaurants, or making non-essential shopping decreased from 20.56% in the first week to 8.61% during the peak of diagnoses.

**Conclusion:** The low proportion of participants that sought medical consultation or stopped leaving home indicates strategies directed to isolate only those symptomatic reach a low proportion of infected patients.

## Introduction

Since March 2020, the SARS-CoV-2 infection became a disease with great concern for the Brazilian public health care system. In the first months, the country was more severely affected by the disease in the regions Southeast and North, with the cities of São Paulo and Manaus having a high death toll^1^. The first case in Joinville, a city in Southern Brazil, was confirmed on March 13, 2020, and up to June 15, there were 24 deaths and 411 diagnoses^2^.

Government responses to COVID-19 have varied worldwide, including enforced social distancing, testing, and contact-tracing, with varying drees of success^3-6^. During the early pandemic phase, the Brazilian Health Ministry’s leadership was unstable, and there was a change of minister of health in April and another in May. Decisions about social distancing were the responsibility of governors and mayors. There was a serious concern with the possibility of pandemic spreading and uncertainties about when and how restrictive public measures could impact each city’s epidemiological situation.

Considering the high uncertainty about the prevalence of exposure to the virus in developing countries, and that comprehensive testing can contribute to a better understanding of the local epidemiological scenario and the planning of public health care interventions, we developed a study to evaluate the serial prevalence of antibodies to SARS-CoV-2 and social distancing practices in representative samples of Joinville, Brazil.

## Methods

### Setting and study design

JoinCOVID was a serial study composed of eight weekly cross-sectional studies that estimated the prevalence of contact with the SARS-CoV-2 virus through serologic tests from June 17 to August 7, 2020, in Joinville, a city in southern Brazil. Joinville is the third most populous city in the three southern states of Brazil. According to the last census, the city had about 598 thousand inhabitants in 2020^7^.

Every week, health care professionals and trained volunteers made telephone calls to households previously randomly selected. A resident was invited to answer a questionnaire that included social distancing practices and symptoms of COVID-19 and perform a serologic test in one of 13 health care centers. The study was approved by a local ethics committee (protocol number 37962620.6.0000.8062).

### The epidemiological scenario and social distancing interventions before the study

The first case of COVID-19 in Brazil was registered on February 25^1^. In a provisional measure taken by the federal judiciary in March, the state’s and city’s governments were considered responsible for social distancing decisions^8^. In Joinville, the first confirmed case of COVID-19 was on March 13^2^. On March 17 and 18, the government of the state of Santa Catarina and the city of Joinville published measures that immediately suspended educational activities, public gatherings, public transportation, and other public services considered “non-essential”. The use of face masks was made mandatory for work and commercial activities^9^. There was no prohibition of people’s circulation in the city (what is often called “lockdown”).

From March 23, the Municipal Health Secretariat started to offer medical evaluations to all inhabitants through telephone calls and messages, for suspicion of COVID-19 or any other medical reason, without out-of-pocket costs. A substantial part of Family Health Units and emergency departments’ activities were directed for caring for people with symptoms of COVID-19, services that are also provided without cost.

### Participants

Multi-stage sampling was used to select participants for the study. We used information provided by the city public institution that supplies water and sewage to define a sampling frame of the households. The institution contains data about 98,3% of the city’s households^10^, being the most reliable database for the researchers.

The households were divided among the strata that correspond to the eight regions of the city and randomized proportionate to the number of inhabitants in each region. A telephone call was performed to each household, and a resident was selected by simple randomization among those older than 18 months. Another qualified resident provided the answers in the case of children or participants with cognitive impairments. A list of substitute households was ordered by zip code to guarantee the proximity of the initially randomized household. In case the phone number was invalid, the call was not answered, or the dwellers did not want to participate, the next household in the list of substitutes was called.

### Variables Collected

A team composed of volunteer healthcare students performed telephone calls, questionnaires, and serological test scheduling, supervised by healthcare professionals from the Health Secretariat. The questionnaire followed a protocol proposed by the World Health Organization^11^, with the addition of some questions to explore aspects related to social distancing practices.

The questionnaire included symptoms, self-reported comorbidities, and practices of social distancing. In case the participant reported coughing, anosmia, coryza, or fever in the last three days, a new interview was scheduled after ten days to avoid exposure to other people during the transportation and serological testing. If the individual continued to present any symptoms in the follow-up call, the participant was excluded from the study and oriented to seek medical evaluation.

The tests were performed in 12 Family Health Units and the Center for COVID-19 Screening of the city.

### Antibody-detecting test

We used the One Step COVID-19 Test (Celer^®^), provided by the Santa Catarina State Department of Health. The test detects the presence of IgG or IgM antibodies to SARS-CoV-2, and possible results are negative or positive. The results were assessed 15 minutes after applying the biological sample, and a trained nurse or doctor issued the report. The test has an estimated sensitivity of 86.43% and specificity of 99.57%. The test’s quality was verified through an evaluation by the National Institute for the Control of Quality in Health of the Oswaldo Cruz Foundation (INCQS/Fiocruz)^12^.

### Statistical analysis

The prevalence of positive cases for COVID-19 was adjusted using the formula^13^:

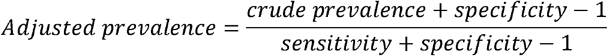

where the crude prevalence was the proportion of positive results, specificity was 99.57%, and sensitivity was 86.43%.

We performed deterministic sensitivity analyses to assess the extent to which the results were modified by changes in the test’s sensitivity or specificity.

The chi-square test was applied to evaluate the difference between participants’ characteristics with positive and negative tests. We considered *p* values below 0.05 to be statistically significant.

The data originating from JoinCOVID were graphically displayed and correlated with the local incidence of diagnosis and deaths by COVID-19 provided by the Health Department of Joinville^2^. Health professionals are required by law to report cases of COVID-19 confirmed by laboratory tests, and the data for correlation included all reported cases. The statistical analyses were conducted using Microsoft Excel 365^®^.

## Results

From June 15 to August 7, 2020, we performed telephone calls for 11 205 households, of which 4561 (40.71%) refused to participate. Among the 6644 (59.29%) persons who answered the questionnaire, 331 (4.98%) had symptoms that might suggest active COVID-19 infection; therefore, their test was rescheduled. Four hundred eighty-eight (7.34%) persons chose not to perform the test after answering the interview. The test was scheduled for 5825 (87.67%) contacts, of which 1422 (24.41%) did not show up for the test. The final sample was 4403 (66.27% of those interviewed, Table 1). The proportion of tests performed per region was very close to the city’s population’s geographical distribution, with the largest deviation occurring in the Southeast region, where 19,18% of Joinville’s population lives, but corresponded to 16,76% of the study’s sample.

**Table 1.** Tests scheduled and performed.

| Week | Period | Tests<br>scheduled | Tests<br>performed | Positive tests |
| --- | --- | --- | --- | --- |
| Week 1 | 15-Jun - 19-Jun | 316 | 245 | 4 (1,63%) |
| Week 2 | 22-Jun - 26-Jun | 582 | 463 | 9 (1,94%) |
| Week 3 | 29-Jun - 03-Jul | 604 | 450 | 13 (2,89%) |
| Week 4 | 06-Jul - 10-Jul | 899 | 671 | 18 (2,68%) |
| Week 5 | 13-Jul - 17-Jul | 1079 | 800 | 28 (3,72%) |
| Week 6 | 20-Jul - 24-Jul | 973 | 714 | 41 (5,74%) |
| Week 7 | 27-Jul - 31-Jul | 796 | 616 | 47 (7,63%) |
| Week 8 | 03-Aug - 07-Aug | 569 | 444 | 53 (11,94%) |
| Total | 15-Jun - 07-Aug | 5.818 | 4.403 | 213 (4,84%) |

The crude prevalence of positive tests varied from 1.63% in the first week (margin of error [ME] 0.05% to 3.22%) to 11.94% in the eighth week (ME 8.92% to 14.95%). After adjusting for the test’s sensitivity and specificity, the estimated prevalence changed to 1.4% (ME 0 to 2.89%) in the first week and 13.38% (ME 10.22% to 16.54%, Figure 1) in the last week. The observed increase in seroconversion presented a high correlation with the city’s count of diagnosis and deaths by COVID-19. We estimate that one in every 5 to 10 of all estimated infections were reported. The deterministic sensitivity analysis indicated that the test’s specificity was 98% or less, the prevalence of positives would be below zero in the first two weeks, which supports the test’s high specificity.

**Figure 1.**
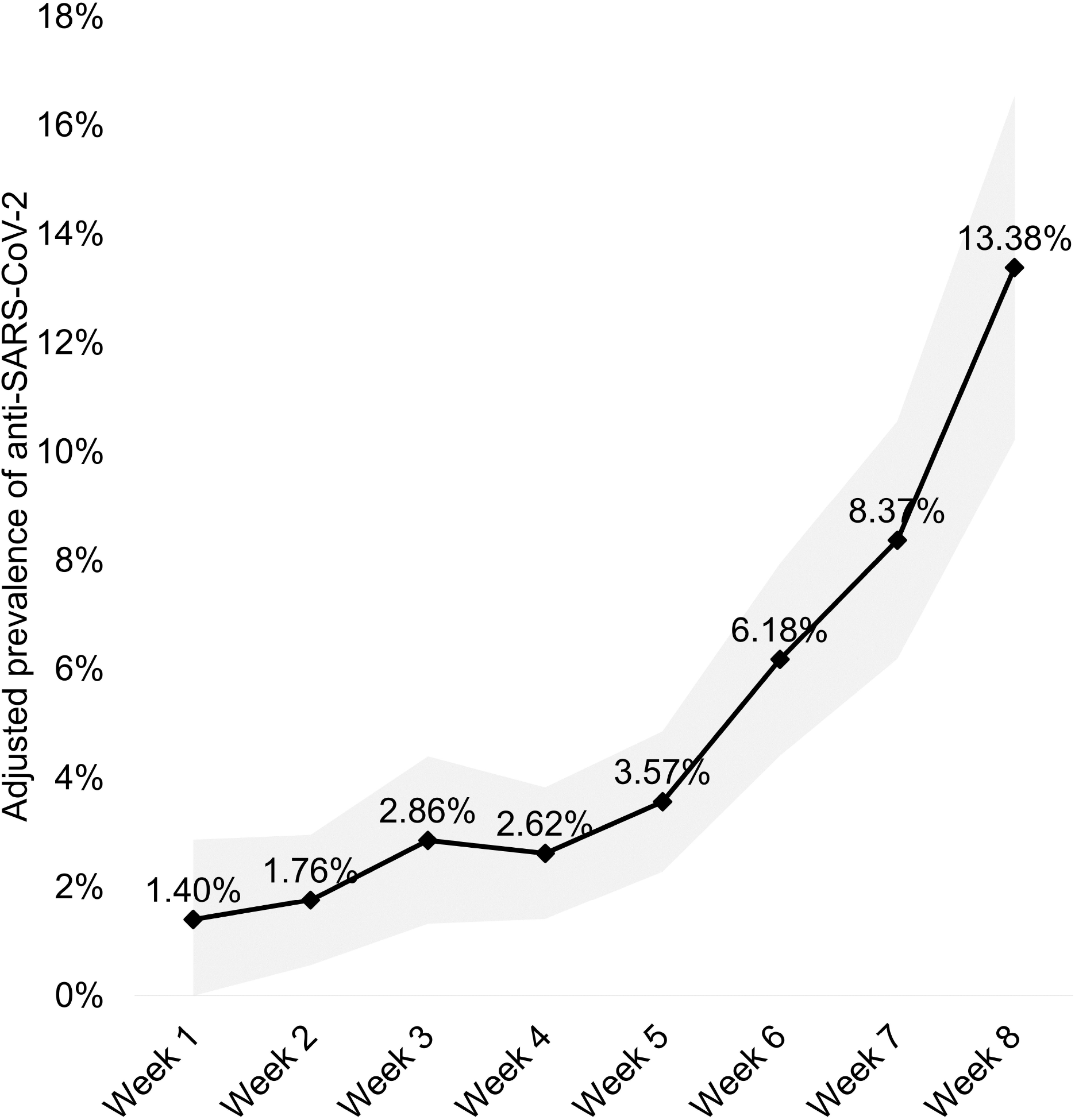
Prevalence of positive tests adjusted for sensitivity and specificity. The points indicate the adjusted prevalence of positives each week, and the shaded area shows the margin of error.

Among the 4403 tests performed, 213 were positive (4.84%), of which 19 (8.89%) are estimated to be false-positive when applying a specificity of 99.57% each week. There was no difference regarding sex (57% of positives vs. 54% of negatives were female, P = 0.3821), the proportion of persons with comorbidities (46% of positives vs. 51% of negatives, P = 0.1241), and the region of residence between positive and negative participants (P = 0.0775). We observed a difference in age between the individuals that tested positive and negative (P = 0.0141), with a higher proportion between ages 30 and 69 (5.3%) and lower between 0 and 29 (2.89%) and older than 70 (2.37%). When the tests were weighted by age, considering the age distribution in Joinville, the total proportion of positives was calculated to be 4.45% (a relative reduction of 8.06%).

Among the 213 participants who tested positive, 64 (30,05%) referred a contact with someone with a confirmed diagnosis of COVID-19. Of all positives, 114 (53.52%) referred fever, coughing, anosmia, or ageusia since March 2020, and 55 (25.82%) did not report any symptoms. Only 37 (17.37%) had sought evaluation with a health care professional due to a symptom. Thirty-three (15.48%) had a confirmed diagnosis of COVID-19 before the questionnaire, of Lwhich 14 (6.57%) were diagnosed using reverse transcription-polymerase chain reaction (RT-PCR) and 19 (8.92%) using antibody-based tests.

Of all individuals who tested positive, 77 (35.81%) referred they were working or studying outside their homes. Of these, only 18 (23.38%) stopped leaving their homes to work or study due to any symptoms since the pandemic. Of the 45 who reported fever, coughing, anosmia or coryza, only 17 (37,77%) stopped leaving their homes to work or study at any moment.

The proportion of individuals that referred leaving home without a mask oscillated between 3.39% (ME 1.81% to 4.98%) in the first week and 1.69% (ME 0.59% to 2.78%) in the final week, reaching a maximum of 3.78% (ME 2.43% to 5.13%) in the third week (Figure 2). The proportion of people that referred going to bars, restaurants, or non-essential shopping was 20.56% (ME 17.02% to 24.1%) in the first week. This proportion started to reduce in the fourth week, reaching 8.61% (ME 6.23% to 10.99%) in the last week.

**Figure 2.**
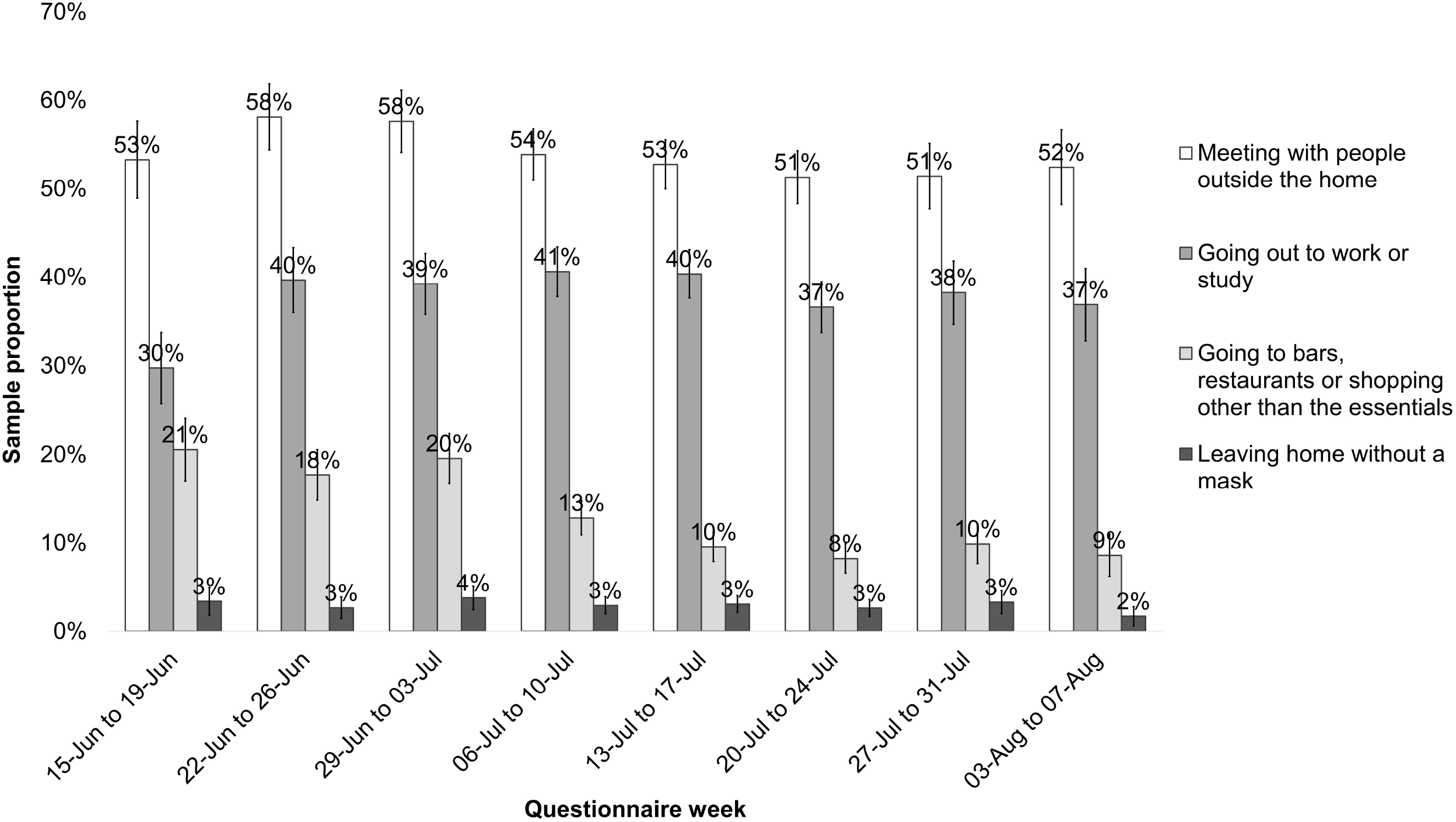
Self-reported social distancing practices. The error bars indicate the margin of error for each activity.

Figure 3 outlines the number of reported COVID-19 diagnoses per day in the city of Joinville. The 7-days moving average number of diagnoses rose from 23 per day on June 17 to 153 on July 1, reaching a peak of 355 on July 28. Figure 4 displays the ICU bed occupancy and the date of governmental decrees with restrictions related to social distancing. The restrictions were imposed with the primary objective of avoiding a demand for ICU beds higher than the city’s supply.

**Figure 3.**
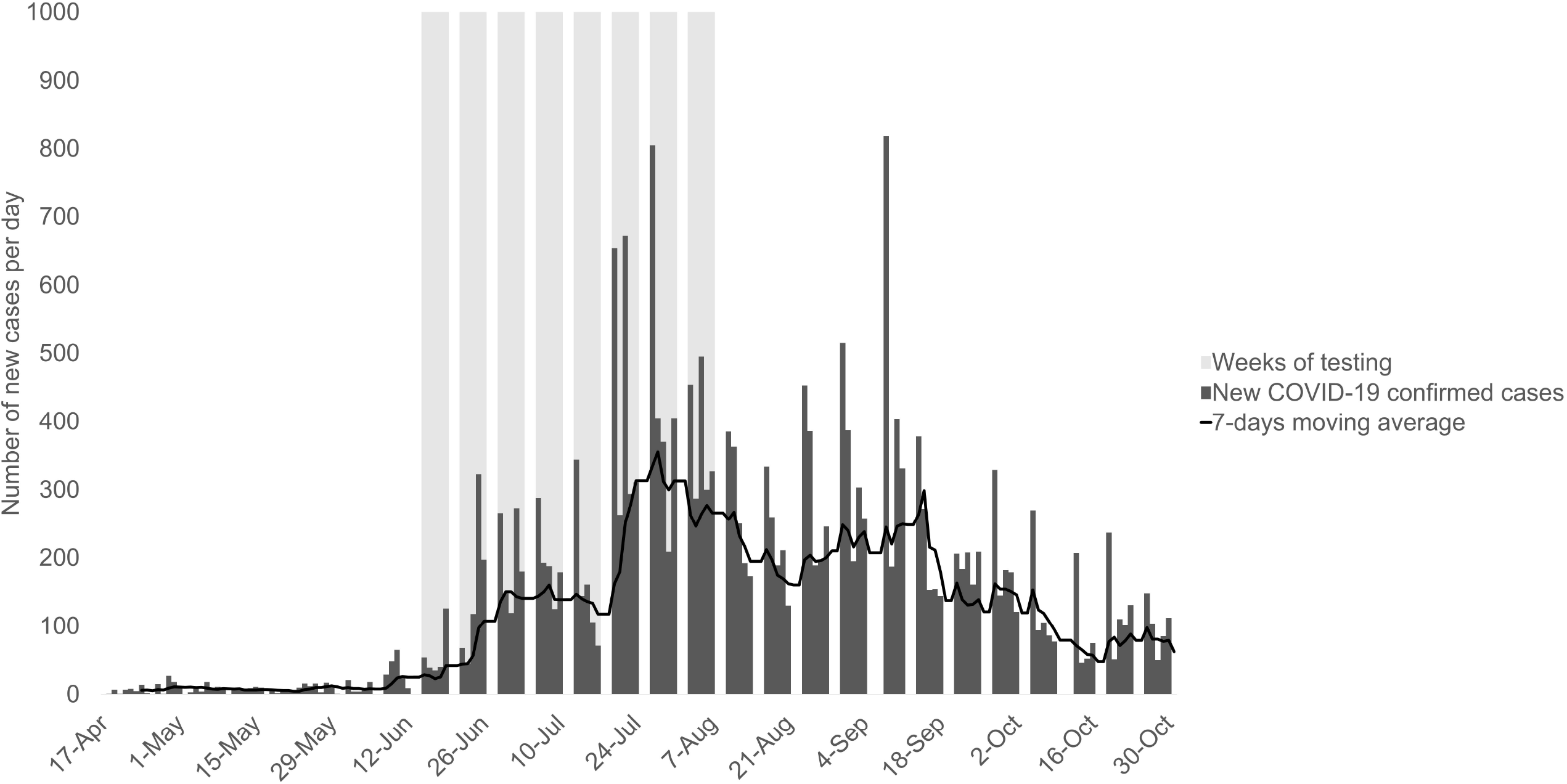
Weeks of testing and the total count of new COVID-19 cases in the city of Joinville. The shade in light gray indicate the eight weeks of testing in JoinCOVID, while the columns in dark gray indicate the number of newly reported diagnoses of COVID-19 in the city, and the black line indicates the 7-days moving average of new COVID-19 diagnoses.

**Figure 4.**
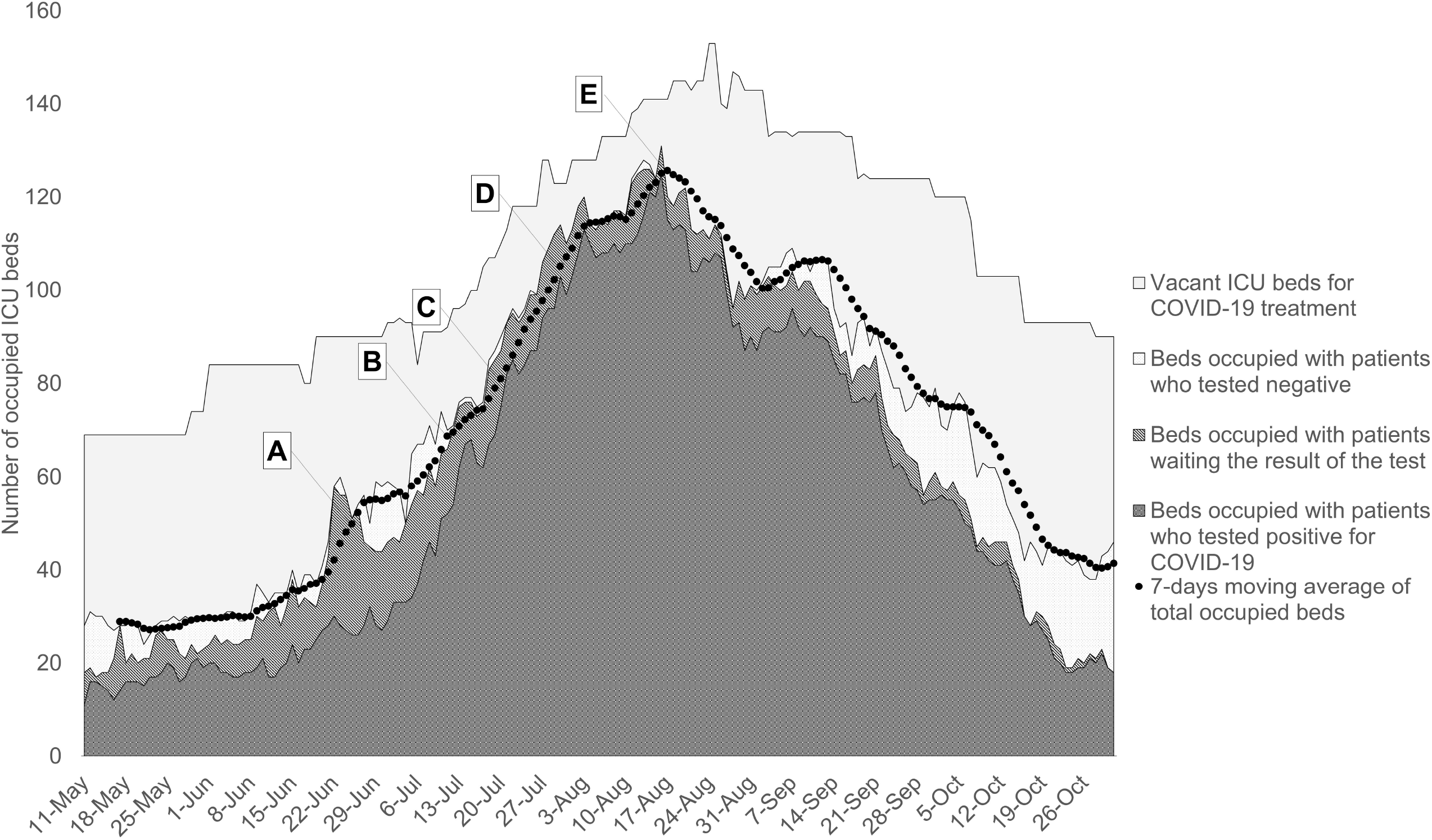
Occupancy of ICU beds, correlated with the governmental decrees issued during the rise in COVID-19 transmission. The letters A to E in the graph indicate the time when each governmental decree was issued. The content of each decree was: A: Mandatory home isolation for people aged 60 and over; suspension of amateur sports and staying in parks and squares; restriction of restaurant occupancy to up to 50%; B: Limitation of the opening hours of bars and restaurants; suspension of all events with public gathering; C: Suspension of municipal public transport; D: Limitation to up to 30% of the maximum service capacity in the following activities: commerce, gyms, religious activities, other face-to-face services (except for health care); entry allowed for up to one person per family in commercial establishments; E: Mandatory home isolation in suspected and confirmed cases of COVID-19, with non-compliance subject to a fine.

## Discussion

The first five weeks indicated a low prevalence of antibodies, with a rapid increase in the last three weeks. This increase accompanied a change in the epidemiological scenario observed through the number of new cases and deaths for COVID-19 and the demand for ICU beds.

A quarter of the participants who tested positive for SARS-CoV-2 did not report COVID-19 symptoms since March 2020, and only 15.49% had a previous laboratory diagnosis. Only 17.37% had sought medical consultation, even though they were available without out-of-pocket payment through various services. Also, less than one in every four participants who tested positive stopped leaving their homes to work or study when they had symptoms suggestive of COVID-19.

Most published studies reported antibodies in less than 4% of the population^14-18^. In some cities, a prevalence between 4 and 10% have been reported^19-21^, and some more heavily affected regions reported a prevalence above 10%^22-24^. One of the most comprehensive studies aimed at obtaining a representative sample of the whole Spanish territory^25^. The study demonstrated that an average of 5% had developed antibodies to COVID-19, with a substantial variation among regions, ranging from 1.4% to 14.4%.

A study conducted in 133 cities in Brazil found a prevalence of 1.6% of positive tests between May 14th and 21st, and 2.8% between June 4 and 7. The study included the city of Joinville, where it tested 250 individuals in each phase, of which none had a positive result. Between 21 and 24 June, the group tested 250 participants, of which 2 (0.8%) had a positive result^26^. Although the study provided a nationwide scenario of the pandemic, it did not provide sufficient information for local decision-making.

A systematic review included 50 155 patients from 41 studies tested with RT-PCR and found that an estimate of 15.6% (confidence interval [CI] 12.1%-23%) were asymptomatic^27^. It is estimated that around 40-45% of COVID-19 infections are transmitted by persons who do not exhibit symptoms^28^. However, to our knowledge, the prevalence of symptomatic patients that continue leaving their homes to work or study is not reported elsewhere.

The current study provides evidence that most people did not stop leaving their homes to work or study, even when they had symptoms of COVID-19. This finding may be explained because a high proportion of infected patients are asymptomatic, have unspecific symptoms, or because patients do not recognize mild symptoms as suggestive of COVID-19. Another significant issue is that workers may be concerned about stopping to work because of mild symptoms, either because they are paid per service or are worried about losing their jobs. Those concerns may be aggravated during an economic crisis. Of note, official estimates report that around 40% of workers in Brazil are informal^29^.

Those findings suggest that public health strategies directed only towards testing and isolating persons with a suspected infection are likely to reach a small portion of potential transmitters. Strategies that target all individuals, such as social distancing, face coverings, and hand hygiene, have more potential to reduce the virus’s transmission since they also encompass asymptomatic or symptomatic residents who do not seek health services.

Our study also provides an example of the weekly use of an epidemiological study for decision-making at the municipal level in a developing country. In a scenario where the mayors have a high degree of responsibility for social distancing practices and the funding, management, and provision of health care resources, JoinCOVID was a useful tool for better understanding the city’s epidemiological scenario.

The information that we still had less than 2% of people exposed to COVID-19 by the end of June, three months after the first case, was critical in providing a picture of the long-term necessity of resources and that the worst period of transmission had not arrived yet. The proportion of positives also helped in estimating a benchmark for the number of recovered, allowing for the creation of more reliable Susceptible-Exposed-Recovered (SIR) models used for the estimation of ICU bed occupancy in the following weeks^30^.

By the end of June and during July, an increase in the transmission rate led to a maximum occupancy of ICU beds, and the region adopted new social distancing measures (Figure 4). Municipal and state public transportation was suspended on July 20. The city also imposed restrictions on activities in restaurants and limited the operation of commerce. The peak in diagnosis occurred on July 28, when JoinCOVID indicated that 8.37% of the sample had developed antibodies. On August 7, when the present study indicated a prevalence of 13.38%, the total number of deaths for COVID-19 was 158^2^. The efficacy of each government response implemented in the city is beyond the present article’s scope. However, having longitudinal population-based data about the prevalence of antibodies and the population’s behavior was crucial in the decision-making process.

### Limitations of this study

A limitation was that the scheduling process was carried out through telephone calls. The process may generate a selection bias in favor of those who have telephone sets and are available to answer the calls. Besides, there was an underrepresentation of residents between 0 and 29 years, and those between 30 and 69 years had a higher representation. On the other hand, an adjustment for age did not substantially change the proportion of residents with a positive test. Besides, the proportion that refused to participate remained similar between regions. The data’s reliability is also strengthened by the high correlation between the total number of positive tests in the city and the prevalence of antibodies in the study.

## Conclusion

In a population-based study of seroprevalence of antibodies to SARS-CoV-2, we found a prevalence ranging from 1.4% in the first week to 13.38% in the eighth week of testing.

Most residents who tested positive did not seek medical attention and continued leaving their homes to work or study. These data indicate a substantial difficulty in controlling the disease’s spread through strategies targeted primarily at diagnosing and isolating residents with suspected disease, justifying more comprehensive measures that increase the social distance between all individuals.

JoinCOVID was a valuable tool to provide a clearer picture of the local epidemiological scenario for decision-makers and to justify the need and timing of decrees related to social distancing practices.

## Data Availability

The anonymized data supporting the findings of the study are available from the corresponding author upon reasonable request.

## Funding

The study was supported through the provision of professionals, working environment, and equipment by the Secretariat of Health of Joinville, the Secretariat of Health of the Santa Catarina State, and Brazil’s Ministry of Health.

## Conflicts of interest

None declared.

## Ethics approval

The study follows all the recommendations from the Declaration of Helsinki and was approved by the corresponding ethics committee by the number 37962620.6.0000.8062.

## Notes

### Competing Interest Statement

The authors have declared no competing interest.

### Author Declarations

This study was approved by the Research Ethics Committee of "Hospital Dona Helena - CEP/HDH", updated on October 2020, approval number CAAE 37962620.6.0000.8062.

